# Neurophysiological biomarkers of post-concussion syndrome: a scoping review

**DOI:** 10.1101/2021.03.25.21254285

**Authors:** Sepehr Mortaheb, Maria Maddalena Filippini, Jean-François Kaux, Jitka Annen, Nicolas Lejeune, Géraldine Martens, Maria Antonia Fuentes Calderón, Steven Laureys, Aurore Thibaut

## Abstract

**Background and objectives:** Post-concussion syndrome (PCS) consists of neurologic and psychological complaints persisting after a mild traumatic brain injury (mTBI). It affects up to 50% of mTBI patients, causes long-term disability and reduces quality of life. The aim of this scoping review was to examine possible uses of different neuroimaging modalities in PCS.

**Methods:** Articles from Pubmed database were screened to extract studies that investigated the relationship between any neuroimaging features and symptoms of PCS. Descriptive statistics were applied to report results.

**Results:** 88 out of 939 papers were included in the final review. 12 examined conventional MRI (42% specificity), 27 diffusion weighted imaging (56% specificity), 25 functional MRI (84% specificity), 10 electro(magneto)encephalography (80% specificity), and 14 examined other techniques (71% specificity).

**Conclusion:** MRI was the most widely used technique, while functional techniques seem to be the most sensitive tools to evaluate PCS. Common patterns associated with symptoms of PCS were a decreased anticorrelation between the default mode network and the task positive network and reduced brain activity in specific areas (most often prefrontal cortex).

**Significance:** Our findings highlight the importance to use functional approaches which demonstrated a functional alternation in brain connectivity and activity in most studies assessing PCS.

**Highlights:**

- post-concussion syndrome causes long term problems for up to 50% of patients after concussion.
- Among different neuroimaging techniques, fMRI and EEG show to be the most sensitive tools for PCS assessment.
- Heterogeneity of axonal injury, symptoms, and populations limits having a specific prognostic criteria for the PCS patients.

## 1. Introduction

Emerging evidence shows that the long-term consequences of concussion – also known as mild traumatic brain injury (mTBI) – can be broad and long-lasting (Hiploylee et al. 2017). Those consequences are known as post-concussion syndrome (PCS); they most frequently comprise complaints as headaches, dizziness, fatigue, irritability, anxiety, insomnia as well as loss of concentration and memory, which can have important socio-economic impact (Junn et al. 2015). Up to 50% of the patients who had a concussion suffer from PCS at 3 months after injury (Voormolen et al. 2019). One study highlighted that 27% of patients who had a concussion could not resume to previous work even after 12 months post-injury; furthermore, among the patients who resumed to previous work, 84% still reported PCS related complaints (Van Der Naalt, Van Zomeren, and Sluiter 1999). These authors also showed that, beside demographics predictors (e.g., age and education) and injury characteristics (e.g., cause and severity of injury), indicators of psychological distress and employment were of influence on work resumption. Proper diagnosis of a concussion is the first step to accurate disease management which can enable further reemployment of subjects and can reduce economic burden on the society. Lack of knowledge (e.g., inaccurate belief that concussion is systematically linked to loss of consciousness) and unfamiliarity with common guidelines in sports’ teams (despite of increasing interest of international federations in establishing new guidelines and regulations in the problem of concussion) are also a serious issue which limits proper diagnosis and appropriate management of patients who suffered from mTBI (McCrea et al. 2004, Kaut et al. 2003).

Heterogeneity of injury and symptoms and current limitations in the sensitivity of imaging and lack of sensitivity of classical biological markers are challenges for the development of diagnostic tools and biomarkers of good and poor recovery. Acute conventional CT (Computed Tomography) findings do not correlate with long-term outcomes such as PCS (Kurča, Sivák, and Kučera 2006; Lee et al. 2008). An extensive meta-analysis suggests that CT should only be performed under certain circumstances, when patients are at risk of severe intracranial injuries and present specific symptoms such as loss of consciousness for more than 5 minutes, declining in neurological status or seizures among others (Easter et al. 2015). Beside CT, other techniques offer alternatives to study brain structural and functional integrity such as MRI (conventional MRI, functional MRI [fMRI], diffusion weighted imaging [DWI], magnetic resonance spectroscopy [MRS]), single-photon emission computed tomography (SPECT), positron emission tomography (PET), magnetoencephalography (MEG), electroencephalography (EEG) or functional near-infrared spectroscopy (fNIRS). Our scoping review aims to address which neuroimaging features of mTBI are relevant to the clinical expression of PCS and could therefore potentially be used as biomarkers for PCS diagnosis and/or prognosis whether cross sectionally or longitudinally.

## 2. Materials and Methods

Search on PubMed database was performed on June 13^th^ 2018 and updated on May 14^th^ 2020 including the following terms: mTBI, mild traumatic brain injury, post concussive syndrome, postconcussion syndrome, post-concussive syndrome, and neuroimaging, magnetic resonance imaging, functional magnetic resonance imaging, diffusion tensor imaging, diffusion spectrum imaging diffusion weighted imaging, susceptibility weighted imaging, diffusion kurtosis imaging, positron emission tomography, computed tomography, single photon emission computed tomography, magnetoencephalography, electroencephalography, near-infrared spectroscopy, functional near-infrared spectroscopy, resting state, functional connectivity.

Inclusion criteria were: 1. articles published in the last 10 years, 2. in humans (adults), 3. on post-concussion syndrome, 4. patients had to be assessed with one of the following techniques: CT, MRI, PET, SPECT, MEG, NIRS or EEG, 5. Published in English.

Exclusion criteria were: 1. articles including less than 12 subjects to exclude case studies and pilot studies (based on the criteria discussed in Julious 2005), and 2. reviews, opinion papers, or case-reports.

Our outcome measure concerned any association between neuroimaging and/or neurophysiological biomarkers and the severity of the symptoms as measured cross sectionally or longitudinally with the following scales (non-exhaustive list): Post-Concussion Symptom Scale, Post-concussive Symptom Questionnaire (PCSQ), PCS-19, PCS-Negative Impression Management (PCS-NIM) Scale, Sport Concussion Assessment Tool (all versions), International Classification of Diseases-10^th^ edition (ICD-10) diagnostic criteria for post-concussion syndrome, Diagnostic and Statistical Manual for mental disorders – IVth edition (DSM-IV), diagnostic criteria for post-concussional disorder, Rivermead Post-Concussion Symptoms Questionnaire (RPQ), British Columbia Post-concussion Symptom Inventory (BC-PSI), Neurobehavioral Symptom Inventory, and Immediate Post-Concussion Assessment and Cognitive Testing (ImPACT).

Two independent blind reviewers (SM, MF) first reviewed the abstracts for inclusion using the Rayyan software (Ouzzani, et al. 2016). In case of disagreement, a third reviewer (AT) was involved in the decision. Descriptive statistics were used to report the results.

## 3. Results

The search yielded 939 articles, from which 851 were excluded and 88 articles were kept for final review (see Fig.1). The imaging techniques used in these 88 studies were conventional MRI (12 papers; 539 participants), DWI (27 papers; 1369 participants), functional MRI (25 papers; 876 participants), EEG/MEG (10 papers; 425 participants), PET (3 papers; 70 participants), SPECT (3 papers; 155 participants), fNIRS (2 papers; 37 participants), MRS (2 papers; 73 participants), and CT (4 papers, 2997 participants; See Fig.2).

**Figure 1.**
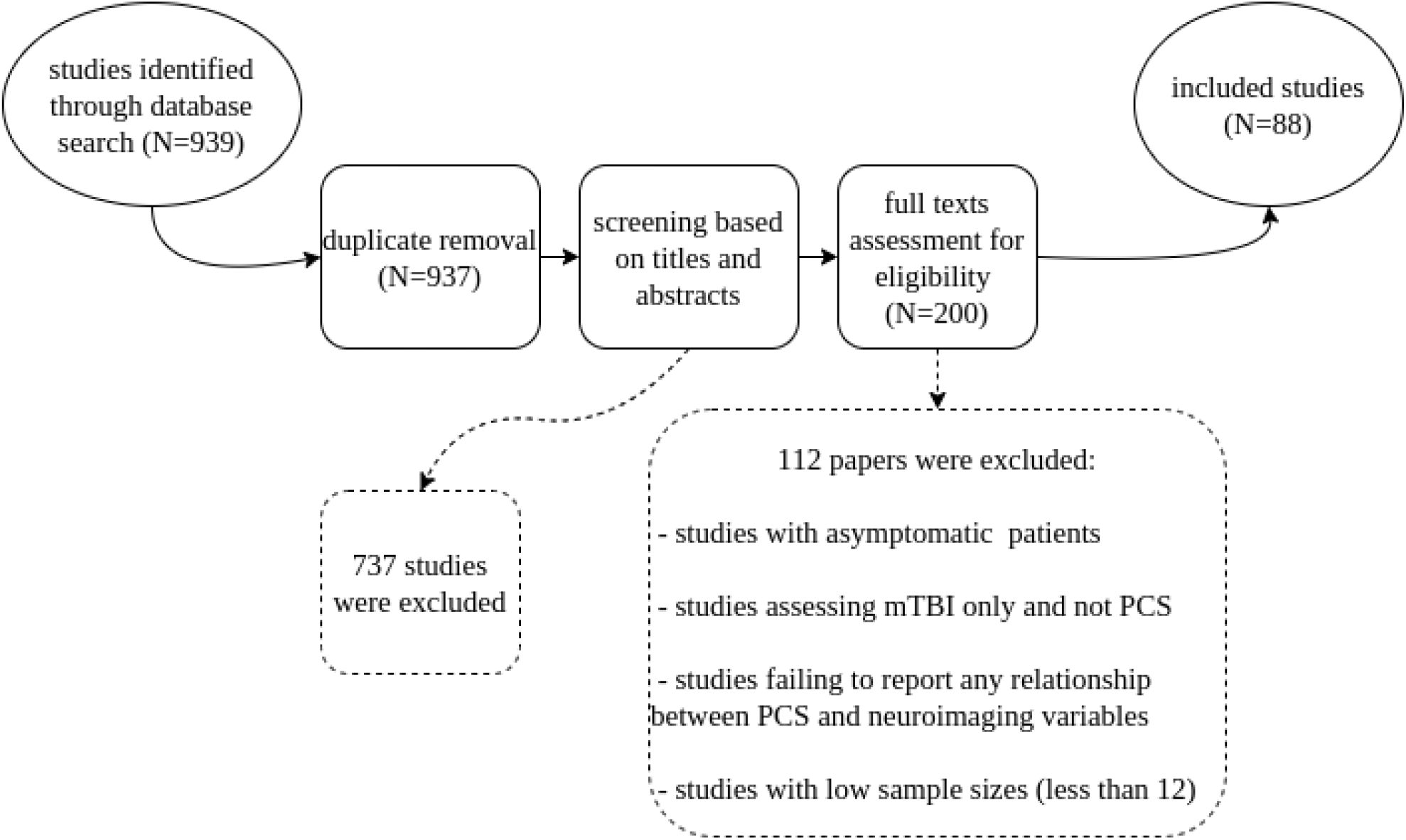
Prisma Flowchart: Systematic search over the Pubmed database yielded 939 studies after which they were screened in the first round based on their abstracts and in the second round based on the main text of the remaining studies. Exclusion criteria were: articles including less than 12 subjects, reviews, opinion papers, and case-reports.

**Figure 2.**
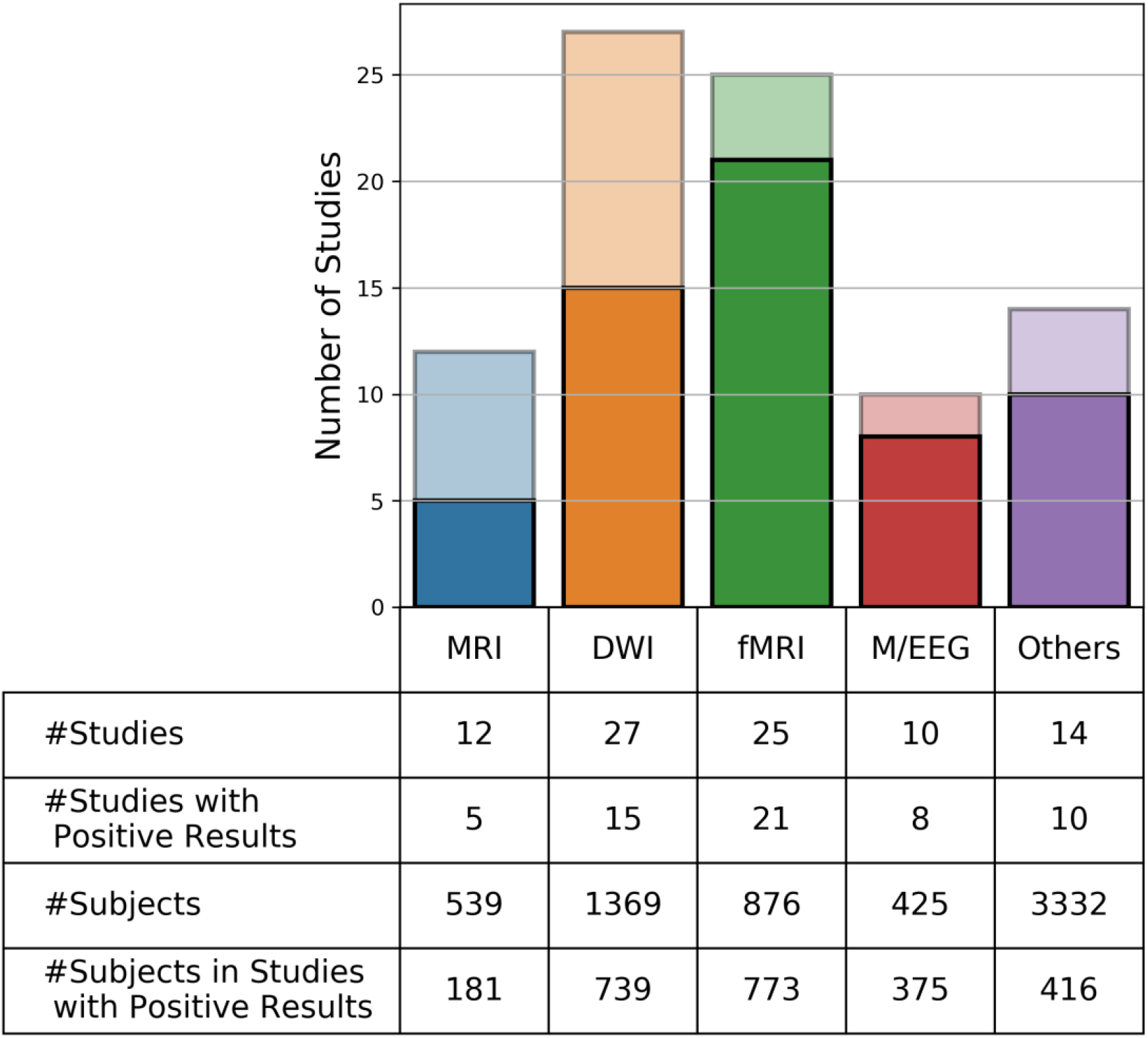
Number of Studies per Imaging Technique: The number of studies we could retrieve per study shows that functional MRI has the highest number of experiments with positive results (i.e. studies which found an association between neuroimaging features and post concussive symptoms). Darker colors show the number of studies with positive results while lighter colors show the studies without significant findings.

We here present results for conventional MRI, DWI, fMRI and EEG/MEG (see Table 1, and for definition of some specific outcome measures mentioned in this table, see Table 2). For the other techniques, only a couple of papers were found which limits the generalizability of the results. The main findings are summarized bellow based on the imaging technique and more extensive results are mentioned in Table 1. In the following, “positive results” means the studies who could find a significant association between at least one neuroimaging marker and PCS.

**Table 1.** Studies included in the final review, sorted by neuroimaging technique. Abbreviations: AD = axial diffusivity; AI = axonal injury; BOLD = blood oxygenation level dependent; DAN = dorsal attentional network; DMN = default mode network; DWI = diffusion weighted imaging; EEG = electroencephalography; ERP = event related potentials; FA = fractional anisotropy; FC = functional connectivity; FLAIR = fluid-attenuated inversion recovery; GM = grey matter; GO = good outcome; ICB = intracranial bleeds; IFO = inferior fronto-occipital fasciculus; ImPACT = immediate postconcussion assessment and cognitive testing; MD = mean diffusivity; MEG = magnetoencephalography; MKT = mean kurtosis tensor; MRI = magnetic resonance imaging; mTBI = mild traumatic brain injury; N = number of patients with mTBI enrolled in the study; na = not available; OFC = orbito-frontal cortex; PCD = post-concussion disorder; PCS = post-concussion syndrome; PCS+ = PCS positive; PCS- = PCS negative; PO = poor outcome; PTSD = post-traumatic stress disorder; RD = radial diffusivity; ROI = region of interest; RPQ = Rivermead Post Concussion Symptoms Questionnaire; RSN = resting state network; SCAT = sport concussion assessment tool; SWI = susceptibility-weighted imaging; TBI = traumatic brain injury; Time related abbreviations (T1 = baseline time point; Tn = follow-up time points; h = hour(s); d = day(s); w = week(s); m = month(s); y = year(s)); vmPFC = ventromedial prefrontal cortex; WM = white matter

| Conventional MRI |  |  |  |  |
| --- | --- | --- | --- | --- |
| Authors (year) | N (PCS) | Time post-injury | Positive results | Main findings |
| (Datta et al. 2009) | 20 (20) | 3-84 m | N | Disturbances of sleep, behaviour and memory and abnormalities in tests for mental speed were more frequent in patients with lesions on MRI, but were not statically significant. |
| (Sigurdardottir et al. 2009) | 40 (25) | 1 y | N | Intracranial pathology did not predict RPQ scores at 12 months post-injury. |
| (Zhou et al. 2013) | 28 (na) | T1: 23 d<br>T2: 1 y | Y | 1 year post-injury, compared to baseline and to healthy controls, mTBI patients showed both global atrophy and atrophy that specifically affected the anterior part of the cingulate white matter (WM) bilaterally, the left cingulate gyrus isthmus WM, and the precuneal grey matter (GM). A negative correlation was found between the cingulate gyrus isthmus volume and clinical symptom scales of PCS at 1 year. |
| (Lannsjö et al. 2013) | 19 (na) | T1: 2-3 d<br>T2: 3-7 m | N | Magnetic resonance pathology was found in one patient at the first examination, but four patients (21%) showed volume loss at the second examination, at which three of them reported less than three symptoms and one of them three symptoms or more, all exhibiting good functional outcome. |
| (Liu et al. 2014) | 63 (36) | 0-7 d | Y | ICBs were detected in more patients and more ICBs were detected on susceptibility weighted imaging (SWI) than T2*-weighted imaging. On SWI more patients from the PCS positive (PCS+) group than the PCS negative (PCS-) group were ICB positive. The numbers of ICBs on SWI were significantly higher in the PCS+ group than the PCS- group. With PCS as the state variable, receiver operating characteristics analysis showed that SWI was superior to T2*WI in PCS prediction. Significant correlation was found between PCS and ICBs number on SWI. Multiple logistic regression analysis showed that ICB number on SWI and superficial location were independent variables predicting occurrence of PCS. |
| (Albaugh et al. 2015) | 16 (na) | na (> 40.6 m) | Y | Cortical thickness in frontal, parietal, and temporal regions was inversely associated with self-reported PCS among collegiate and preparatory school male ice hockey players. |
| (Killgore et al. 2016) | 26 (na) | 2 w -1 y | Y | Grey matter volume within the vmPFC and the fusiform gyrus was positively correlated with some tests of executive functioning (design fluency, sustained visual psychomotor vigilance test). Volume within the vmPFC negatively correlated with symptoms of anxiety. |
| (Clark et al. 2016) | 46 (46) | 6-121 m | N | Neither deep nor periventricular white matter hyperintensities detected on FLAIR (Fluid-Attenuated Inversion Recovery) images significantly predicted post-concussive symptoms. |
| (Hellstrøm et al. 2017) | 62 (na) | T1: 30.42 d<br>T2: 1 y | N | No significant differences in PCS symptoms were observed between mTBI patients with and without intracranial pathology on CT or on structural MRI, or both at 12 months post-injury. 12 months after mTBI, significant within-group volume changes were detected in several brain areas for both the uncomplicated mTBI group and the complicated mTBI group. No associations between volumetric variables and post-concussive symptoms or global functioning were found. |
| (de Haan et al. 2017) | 127 (na) | 24 (14) w | N | Both the mean number of complaints and the presence of specific complaints of memory and concentration were not different between groups of patients with and without MRI abnormalities. There was no significant correlation between the number of microhaemorrhages and the number of post-traumatic complaints. The presence of memory and concentration complaints also was not significantly correlated with the number of microhaemorrhages. An unfavourable functional outcome in the chronic post-traumatic phase was associated with the presence and number of microhaemorrhages in the temporal cortical area. |
| (Chai et al. 2017) | 48 (na) | na | Y | In a group of mTBI patients with MRI signs of axonal injury (AI), a negative correlation between a measure of cerebral venous oxygen saturation in the straight sinus and the RPQ scores was observed, but no significant correlation was found in the non-AI group. There were no correlations between measures of cerebral venous oxygen saturation in other cerebral veins (the right and left cortical veins; septal veins; thalamostriate veins; internal cerebral veins; basal veins) and RPQ scores in the AI or the non-AI groups. |
| (Yoo et al. 2019) | 44 (44) | 4 (2-37.5) m | N | No significant correlations were noted between PCS symptom score and dynamic contrast-enhanced MR imaging parameters for the quantitative analysis of blood-brain barrier disruption. |

Diffusion Weighted Imaging
| Authors (year) | N (PCS) | Time post-injury | Positive results | Main findings |
| --- | --- | --- | --- | --- |
| (Smits et al. 2011) | 19 (na) | 18-40 d | Y | When comparing patients with controls, no differences were found in mean diffusivity (MD). One area of decreased fractional anisotropy (FA) was found in patients compared with controls in the right temporal lobe subcortical fibres of the inferior fronto-occipital fasciculus (IFO). Significant increase of MD associated with the severity of post-concussive symptoms was seen in the left IFO and inferior longitudinal fasciculus, as well as in the superior longitudinal fasciculus. A significant reduction of FA in association with the severity of post-concussive symptoms was seen in the right uncinate and IFO, the posterior limb of the internal capsule bilaterally, the splenium of the corpus callosum on the right side, as well as in peripheral white matter consisting of fibres originating from the corpus callosum in the left parietal and right frontal lobes. |
| (Messé et al. 2011) | 23 (12) | T1: 7-28 d<br>T2: 3-4 m | Y | Higher MD values were observed in poor outcome (PO – persistent PCS at 3 months post injury) patients compared with both controls and good outcome (GO – no persistent PCS at 3 months post-injury) patients, in the forceps major and minor of the corpus callosum, the IFO bilaterally, and the inferior longitudinal fasciculus bilaterally. PO patients showed higher MD values than controls in the superior longitudinal fasciculus and the corticospinal tract bilaterally, and the left anterior thalamic radiation. An index of MD was significantly higher for PO patients than for controls and GO patients whereas they did not differ between GO patients and controls. |
| (Messé et al. 2012) | 53 (22) | T1: 8-21 d<br>T2: 6 m | Y | mTBI patients presented decreased FA values in association, commissural and projection white matter fiber tracts and these changes evolved over time. When comparing mTBI patients on the basis of their complaints, PCS+ patients had greater and wider impaired brain regions (i.e. with diffusion measures significantly different) than PCS- patients, at the subacute and late phases. Moreover, significant difference was found between PCS+ and PCS- patients in MD and axial diffusivity (AD) values primarily in the corpus callosum, the longitudinal fasciculus and the internal capsules. |
| (Lange et al. 2012) | 60 (21) | 47 (6,3) d | N | There were no significant differences between mTBI and trauma control groups on all diffusion and anisotropy measures. In the mTBI sample, there were no significant differences on all diffusion and anisotropy measures between those who did and did not meet the criteria for PCS. There was, however, a small-medium effect size for FA in the genu and body of the corpus callosum. For these measures, there was a non-significant trend for lower FA in the post-concussion disorder present (PCD-present) group compared with the post-concussion absent (PCD-absent) group. |
| (Fakhran, Yaeger, and Alhilali 2013) | 64 (na) | 1-486 d | Y | Quantitative comparison for tract based spatial statistics analysis between patients with mTBI and control subjects showed widespread significant differences in FA. In no regions did patients with mTBI have significantly higher FA values than those in control subjects. Total concussion symptom scores correlated positively with FA values at the GM-WM junction, most prominently at regions of geometric inflection and in the primary and association auditory cortices. There were no regions where FA values negatively correlated with total concussion symptom scores. |
| (Yeh et al. 2014) | 29 (na) | 334.5-503.8 d | Y | Scores on a self-report of post-concussion symptoms were significantly associated with low FA mainly in the regions of the junction of ventral striatum, genu of internal capsule, optic chiasma near the level of midbrain and the ventral (hippocampal) part of the left cingulum bundle and the left thalamus. Looking at blast and non-blast traumatic brain injury (TBI) groups separately, greater PCS in the non-blast subjects were associated with lower FA in the unilateral anterior-posteriorly oriented tracts, such as anterior and posterior parts of dorsal cingulum bundle, and commissural fibers such as splenium of corpus callosum. |
| (Ilvesmäki et al. 2014) | 43 (32) | 48.1 (45.4) h | N | Acute mTBI was not associated with diffusion and anisotropy abnormalities detectable with tract-based spatial statistics. |
| (Wäljas et al. 2014) | 37 (11) | 16-60 d | N | mTBI group had a significantly larger number of FA values compared to the healthy controls. mTBI patients with multifocal WM changes did not show evidence of worse symptoms, cognitive impairment, or slower return to work compared to mTBI patients with broadly normal WM. |
| (Petrie et al. 2014) | 34 (na) | 1.2–7.1 y | N | Compared with non-blast veterans, the blast-mTBI veterans endorsed more frequent and severe PCS. Within the blast-mTBI veteran group, correlation analyses showed no statistically significant associations (after corrections for multiple comparisons) between FA or macromolecular proton fraction values and any of the behavioural or neurological assessment measures. |
| (Panenka et al. 2015) | 62 (na) | 42-56 d | N | No significant differences were observed between complicated (i.e. with intracranial abnormalities assessed with neuroimaging) and uncomplicated mTBI groups on post-concussion symptom reporting, although the complicated group showed microstructural differences on DWI, particularly in the corpus callosum and frontal areas. |
| (Davenport, Lim, and Sponheim 2015) | 125 (na) | 2-5 y | N | No WM integrity measures were significantly associated with PCS scores. |
| (Lange et al. 2015) | 72 (20) | 46,70 (5,5) d | Y | For the diffusion and anisotropy measures, there were no significant differences in FA, AD, radial diffusivity (RD) or MD when comparing the PCS-present and PCS-absent groups. However, there were significant differences in MD and RD when comparing the PCS-present and trauma control groups. RD was significantly increased in the PCS-present group in widespread areas including the genu, body, and splenium of the corpus callosum; bilateral anterior, superior, and posterior corona radiata; bilateral superior longitudinal fasciculus; bilateral cingulum; bilateral posterior thalamic radiations; bilateral anterior limb of the internal capsule; and bilateral external capsule. MD values were also significantly increased in the PCS-present group in the genu and body of the corpus callosum, bilateral superior corona radiata, right anterior corona radiata, right anterior and posterior limb of the internal capsule, and the right superior longitudinal fasciculus. Within the mTBI group, WM changes were not a significant predictor of PCS. |
| (Alhilali et al. 2015) | 45 (na) | 0-506 d | Y | Patients with mTBI who had developed depression had significantly lower FA in the region of the right nucleus accumbens, anterior limb of the internal capsule, and superior longitudinal fasciculus. No significant correlation was found between mean FA in nucleus accumbens region of interest (ROI) and Symptom Severity Score. |
| (Wäljas et al. 2015) | 126 (124) | 8-38 d | N | Microstructural WM findings were not significantly associated with greater post-concussion symptom reporting. |
| (Delano-Wood et al. 2015) | 38 (na) | 16-259 m | Y | Lower FA of certain WM tracts in the brainstem —especially the pontine tegmentum—was significantly associated with increased PCS symptoms (i.e., more severe vestibular symptoms, poorer physical functioning, and greater levels of fatigue), even after adjusting for post-traumatic stress disorder (PTSD) symptoms. |
| (Miller et al. 2016) | 53 (na) | 50 m | Y | Patients who suffered a more severe mTBI (i.e. with loss of consciousness) had a greater number of clusters with reduced FA compared to no-TBI sub- jects. mTBI-associated white matters abnormalities were spatially heterogeneous. Physical post-concussion symptom severity was associated with the number of clusters with reduced FA, even after correcting for PTSD. The number of clusters with reduced FA was a significant mediator between mTBI and loss of consciousness and physical post-concussion symptom severity. |
| (Meier et al. 2016) | 40 (na) | T1: 1.64 d<br>T2: 8.33 d<br>T3: 32.15 d | N | Concussed athletes exhibited increased FA in several WM tracts at each visit post-concussion with no longitudinal evidence of recovery. Correlation between behavioural outcomes and metrics of increased FA was not significant. |
| (Lancaster et al. 2016) | 26 (na) | T1: 14-24 h<br>T2: 8 d | Y | Within 24 hours after injury, the concussed group exhibited a widespread decrease in MD, increased axial kurtosis and, to a lesser extent, decreased AD and RD compared with control subjects. There were also notable associations between diffusion metrics and symptom severity measured by the SCAT-3 at both time points in the total sample. Higher SCAT-3 symptom severity scores correlated significantly with decreased mean diffusion, AD and RD and increased axial kurtosis metrics obtained at 24 hours in the total sample. Day 8 SCAT-3 symptom severity scores still correlated with diffusion measures at day 8 in the total sample, with increased symptom burden related to reduced diffusion and increased axial kurtosis. SCAT-3 scores at 24 hours correlated significantly with diffusion metrics on day 8. More specifically, greater symptom severity on the SCAT-3 at 24 hours predicted lower diffusion and higher axial kurtosis at 8 days after injury. |
| (Astafiev et al. 2016) | 20 (na) | na | N | Correlations of FA values in the ROI near the left optic radiation with a clinical PCS scale was not significant, although there was a trend for a positive correlation. |
| (Thomas et al. 2017) | 20 (20) | T1: 46.5 (22) h<br>T2: 1 w | Y | Significant correlations were found between PCS symptom scores at one week follow up and (i) the FA of left uncinate fasciculus at time period 1, (ii) longitudinal changes in the FA of corona radiata, and (iii) absolute values of longitudinal changes in FA in 6 other ROIs. |
| (Yeh et al. 2017) | 202 (202) | 576 (238.9) d | Y | WM microstructural injury in chronic blast mTBI patients was observed over the frontal fiber tracts, that is, the front-limbic projection fibers (cingulum bundle, uncinate fasciculus), the fronto-parieto-temporal association fibers (superior longitudinal fasciculus), and the fronto-striatal pathways (anterior thalamic radiation). PCS and PTSD symptoms as well as neuropsychological function were associated with low FA in the major nodes of compromised neurocircuitry. |
| (Næss-Schmidt et al. 2017) | 27 (na) | T1: 14 d<br>T2: 3 m | N | Mean kurtosis tensor (MKT) decreased significantly from baseline to follow-up in the thalamus. Compared to healthy controls, thalamic MKT values were also significantly elevated at baseline. Post-hoc analysis of the splenium in the corpus callosum revealed a significant decrease in FA between baseline and follow-up. No correlation was observed between change in specific symptoms and change in MKT of the thalamus or FA in the splenium. |
| (Rangaprakash et al. 2017) | 42 (42) | na | Y | DTI results revealed greater diversity in structural connectivity between hippocampus and striatum, with WM fibers connecting these two ROIs being more diffuse (implying less integrity) in the PCS-PTSD group compared to the PTSD-only and control groups. The profile was similar in the control and PTSD-only groups, suggesting a strong structural basis for PCS. |
| (Klimova et al. 2018) | 40 (na) | 126.6 (121.4) | N | In a group of mTBI patients compared to subjects with PTSD without prior head trauma, stepwise regression analysis was conducted to identify the relative associations of FA changes and PTSD severity to PCS in those WM tracts that showed significant differences between mTBI and PTSD group. Postconcussive symptoms were largely explained by PTSD severity rather than by changes in brain WM. |
| (Lancaster et al. 2018) | 20 (na) | T1: 19.69 (14–24) h<br>T2: 8.38 (7–11) d<br>T3: 168.56 (151–204) d | Y | Mean and axial diffusivity in several major WM pathways (corpus callosum, limbic association fibers, long association fibers, and projection fibers) were significantly lower in concussed athletes at 6 months postinjury compared to nonconcussed athlete controls. More postconcussive symptoms reported within 24 hr of injury were associated with lower diffusion measures at 6 months postinjury, suggesting a dose dependent relationship whereby more symptomatic concussions may lead to greater long-term differences in this diffusion metric. |
| (Murdaugh et al. 2018) | 16 (na) | T1: 0-7 d<br>T2: 21-28 d | N | Increased isotropic value of the diffusion orientation density function along a segment of the cortico-spinal tract was observed in concussed participants versus controls at the acute phase post-injury. No significant correlations were found between total symptoms scores and diffusion metrics. |
| (Yin et al. 2019) | 33 (na) | T1: 2 (0-5) d<br>T2: 37 (27-35) d<br>T3: 104 (85-105) d | Y | Persistent loss of integrity was observed in the right anterior corona radiata, forceps major and body of corpus callosum beyond 3 months post-injury. Reduced FA values in the forceps major were negatively correlated with baseline PCS symptoms score. |

| Functional MRI |  |  |  |  |
| --- | --- | --- | --- | --- |
| Authors (year) | N (PCS) | Time post-injury | Positive Results | Main findings |
| (Smits et al. 2009) | 21 (21) | 30 d | Y | Severity of PCS is related to brain activation increase during verbal working memory and selective attention processing, and to activation outside the working memory network in high working memory load tasks. |
| (Pardini et al. 2010) | 16 (na) | 7 d | Y | PCS symptom severity is related to increased BOLD (blood oxygen level dependent) response within premotor cortex, posterior parietal cortex. Severity of PCS is related to more cognitive resources needed to perform the same task. |
| (Tang et al. 2011) | 24 (na) | 22 d | Y | Severity of PCS is related to reduced degree of symmetry of the thalamic resting state networks (RSN). |
| (Gosselin et al. 2011) | 14 (na) | 5.7 m | Y | Higher PCS scores is related to lower BOLD signal changes in the right mid dorsolateral prefrontal cortex during a visual externally ordered working memory task, and to higher BOLD signal changes in the left parietal region. |
| (Stevens et al. 2012) | 30 (na) | 60.9 (35.77) d | Y | More severe PCS symptoms are related to less connectivity in the primary visual network, cingulo-opercular network, right frontoparietal network and default mode network (DMN). Greater number of complaints is related to diminished anterior cingulate connectivity in five discrete RSNs: an emotional network, two higher-order processing networks, the ventromedial portion of the DMN, and the primary visual processing circuit. In the primary visual processing circuit, greater PSC scores is related to lesser functional connectivity (FC) of bilateral lingual gyri. Higher PCS scores are related to enhanced connectivity of the left frontoparietal network and mediofrontal network. |
| (Terry et al. 2012) | 20 (9) | 596.17 d | N | No main effect of group comparing the functional activation of symptomatic concussed athletes to asymptomatic concussed athletes during all of the functional tasks.<br>No differences in activation comparing the symptomatic sub-group to the non-concussed group. |
| (Zhou et al. 2012) | 26 (na) | 22 d | Y | Functional connectivity of anterior medial prefrontal cortex is related to anxiety, depression, fatigue, and post-concussive symptoms. |
| (Messé et al. 2013) | 55 (17) | T1: 1-3 w<br>T2: 6 m | Y | PCS+ vs PCS- and healthy controls: graph topological properties of RSNs increased in temporal regions for subacute patients and decreased in frontal regions for chronic patients. |
| (Mutch et al. 2014) | 12 (12) | na | Y | Alterations in regional cerebrovascular flow to provocative CO2 challenge in concussion patients, not in healthy controls. |
| (Sours, Chen, et al. 2015) | 32 (15) | T1: 6 d<br>T2: 6 m | Y | Overall reduction in connectivity in 0.125-0.250 Hz frequency range in PCS+ vs PCS-. Reduced strength of connectivity in PCS+. |
| (Sours, Zhuo, et al. 2015) | 28 (12) | T1: 7 d<br>T2: 1 m<br>T3: 6 m | N | No difference between PCS- and PCS+ in DMN or temporo-parietal network cerebral blood flow. |
| (Sours, George, et al. 2015) | 77 (na) | 6 d | Y | Increased spatial extent of FC between the thalamus and cortical areas. Increased FC between pre motor sub-thalamic ROI and posterior cingulate cortex. Increased FC between prefrontal cortex sub-thalamic ROI and dorsal anterior cingulate cortex and bilateral medial temporal lobe. Increased connectivity between the 7 subthalamic ROIs and the nodes of the DMN. |
| (Iraji et al. 2015) | 12 (na) | 0.54 d | N | Standardised assessment of concussion scores were not correlated with FC. |
| (Koski et al. 2015) | 12 (12) | T1 : na<br>T2 : 2 w<br>T3: 3 m | Y | Decline in PCS severity and increase in working memory task performance after transcranial magnetic stimulation (not present at 3m follow-up), correlated with a greater activation in dorsolateral prefrontal cortex and greater reduction in activity of the anterior cingulate cortex. |
| (Astafiev et al. 2016) | 20 (na) | na | Y | Larger task-evoked BOLD activity in middle temporal and lateral occipital visual areas in mTBI and PTSD symptoms.<br>Different parts of the WM have reduced FC with the middle temporal and lateral occipital visual areas ROI in mTBI patients. |
| (Van der Horn et al. 2016) | 52 (32) | 30.42 d | Y | Lower activation in the medial prefrontal cortex during high working memory load. Stronger deactivation of the DMN and lower functional connectivity between DMN and fronto-executive network in PCS-. |
| (Meier et al. 2016) | 43 (na) | T1: 1.74 d<br>T2: 1 w<br>T3: 1 m | N | No significant correlations between behavior and connectivity measures at T1, T2, or T3. |
| (Banks et al. 2016) | 13 (na) | T1: 42 d<br>T2: 4 m | Y | Poorer performance in neuropsychological tests is related to increased thalamus and thalamus-DMN FC and decreased in thalamus-dorsal attention network (DAN) and thalamus-frontoparietal connectivity. At 4 months follow up, improvement in neuropsychological test measure and PCS symptoms are related to the trend to normalisation of FC patterns (except for thalamus-DMN functional connectivity). PCS symptoms are related to changes in thalamus-dorsal attentional network FC; as thalamus-DAN FC normalized, pain and post-concussive symptoms improved. |
| (Palacios et al. 2017) | 75 (na) | T1: 11.2 d<br>T2: 6 m | Y | Negative correlations between semi-acute connectivity and RPQ score at 6 months were found within the posterior regions of several networks (visual network; occipito-cerebellar network; dorsal visual stream; posterior default mode network) only in the CT/MRI PCS- group. Decreased connectivity is related to PCS symptoms severity. |
| (Zhou 2017) | 30 (na) | 22 d | Y | Lower relative betweenness centrality in mTBI and higher clustering coefficient. Higher thalamocortical connectivity in mTBI. Higher clustering coefficient is related to reduced PCS score. |
| (Murdaugh et al. 2018) | 28(16) | T1: 7 d<br>T2: 21 d | Y | Resting state hyperconnectivity within posterior brain regions. Hypoconnectivity in anterior areas. Positive correlation between performance in IMPACT and resting state network connectivity at both time points. |
| (Chong et al. 2019) | 15(15) | T1: 1 m<br>T2: 5 m | Y | Weaker homotopic functional connectivity of pain processing regions in PCS+ patients compared to healthy controls. In 5 month followup, symptom recovery was correlated with homotopic functional connectivity strengthening of pain processing regions as well as primary somatosensory area. |
| (Hou et al. 2019) | 47(24) | 7 d | Y | Greater minimum spanning tree and average shortest path of static functional connectivity networks in PCS+ shows their costly and inefficient within network communication.<br>PCS+ patients have slower dynamic changes and mainly tend to keep a functional configuration which is characterized by disruption of small world topology. |
| (Kaushal et al. 2019) | 122 (62) | T1: 1.35 d<br>T2: 8.2 d<br>T3: 15.42 d<br>T4: 45.56 d | Y | Higher network connectivity in the PCS+ group at 8 days. No significant differences with PCS- at the other time points. Functional connectivity is significantly altered 8 days after PCS when symptoms are still reported. Recovery of resting state FC abnormalities follows a trajectory similar to clinical recovery. |
| (Manning et al. 2019) | 52(21) | T1: 3 d<br>T2: 3 m<br>T3: 6 m | Y | In a structure-function study, alterations of diffusion measures in the fornix and DMN and executive RSN hyperconnectivity was correlated with SCAT3 symptom score. |

| EEG/MEG |  |  |  |  |
| --- | --- | --- | --- | --- |
| Authors (year) | N (PCS) | Time post-injury | Positive Results | Main findings |
| (Gosselin et al. 2011) | 14 (na) | 5.7 m | N | When the mTBI and control subjects were pooled, correlations between N350 amplitude change (difference between the working memory and control tasks), and BOLD signal change in the right mid-dorsolateral prefrontal cortex were found. Similar results were obtained using PCS as a covariate ($r = -0.51$, $p < 0.01$), showing that greater change in the N350 event related potential (ERP) component was associated with greater BOLD signal change. No significant correlation between functional MRI and ERP variables was found when the mTBI group was analyzed separately, and no correlation was found with other demographic or clinical variables. |
| (Gosselin et al. 2012) | 44 (na) | 7.6 m | Y | A group effect was observed for the N350 amplitude in the decision phase for the working memory condition, where patients with mTBI showed smaller amplitude in comparison with controls. A group effect was observed for the P300 amplitude in the decision phase for the working memory condition. As observed with the N350 wave, patients with mTBI had smaller amplitudes for the P300 wave when compared with controls. |
| (Larson, Clayson, and Farrer 2012) | 36 (na) | 212.92 d | N | No correlation between RPQ scores and electrophysiological measures. |
| (Huang et al. 2014) | 84 (84) | 264.63 d | Y | MEG source magnitude is related to PCS scores.<br>- mTBI:<br>1) personality change symptoms (e.g., social problems) is related to MEG slow-wave generation in bilateral orbitofrontal cortex (OFC) and vmPFC.<br>2) Trouble concentrating and affective lability (quickly-changing emotions) |
|  |  |  |  | symptoms were associated with slow-wave generation in right OFC.<br>3) Blurred vision or other visual difficulties symptoms is related to slow-wave generation in right fusiform gurus.<br>- non-blast mTBI.<br>4) Depression symptoms is related to slow-wave generation in anterior cingulate cortex.<br>- Combining the blast and non-blast mTBI:<br>5) MEG source magnitude from the right OFC is related to symptoms of trouble concentrating. |
| (Robb Swan et al. 2015) | 31 (31) | 97.4 d | Y | Slow-wave activity on MEG is related to patterns of cognitive functioning in cortical areas, consistent with cognitive impairments on exams. |
| (Vakorin et al. 2016) | 20 (na) | 32 d | Y | Classification of mTBI and healthy controls:<br>1) Total accuracy have a local maximum around 8-13 Hz ($\alpha$ rhythms).<br>2) With an accuracy of 80% inter-regional resting state phase synchrony in the $\alpha$ band carries the most discriminative information for inferring the presence or absence of mTBI within a single individual.<br>The confidence of classifying a subject as mTBI is related to self-reported severity scores and symptoms. |
| (Huang et al. 2017) | 26 (26) | 507.35 d | Y | Well-matched military control group vs blast mTBI group:<br>1) Increased ROI global FC in the prefrontal cortex, medial temporal lobe, putamen, and cerebellum in mTBI.<br>2) FC in right frontal pole was decreased in blast mTBI.<br>3) Abnormal FC beta and low-frequency bands, with some in the gamma band; however, alpha-band intrinsic FC was preserved in mTBI. |
| (Reches et al. 2017) | 80 (na) | T1: 2-10 d<br>T2a: 7 (2) d after T1<br>T2b: 17.2 d after T1 | Y | Among patient older than 16 years, a significant difference in brain network activation score was obtained between subjects in the concussion and control groups at the first visit. |
| (Lewine et al. 2019) | 71 (71) | > 5m | Y | Increase of relative theta power and decrease of relative alpha power (whole brain). Decrease of beta inter-hemispheric coherence. |
| (Ruiter et al. 2019) | 19 (19) | 28.11 y | Y | Reduction of P300, N2b, N1 and MMN amplitude in patients compared to healthy controls. |

**Table 2.**
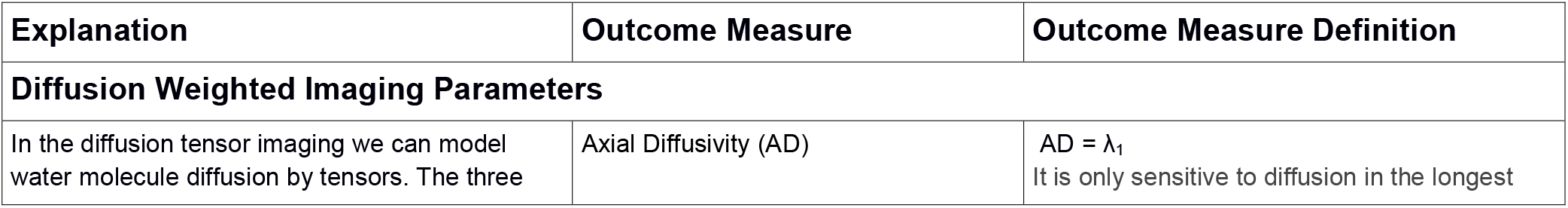
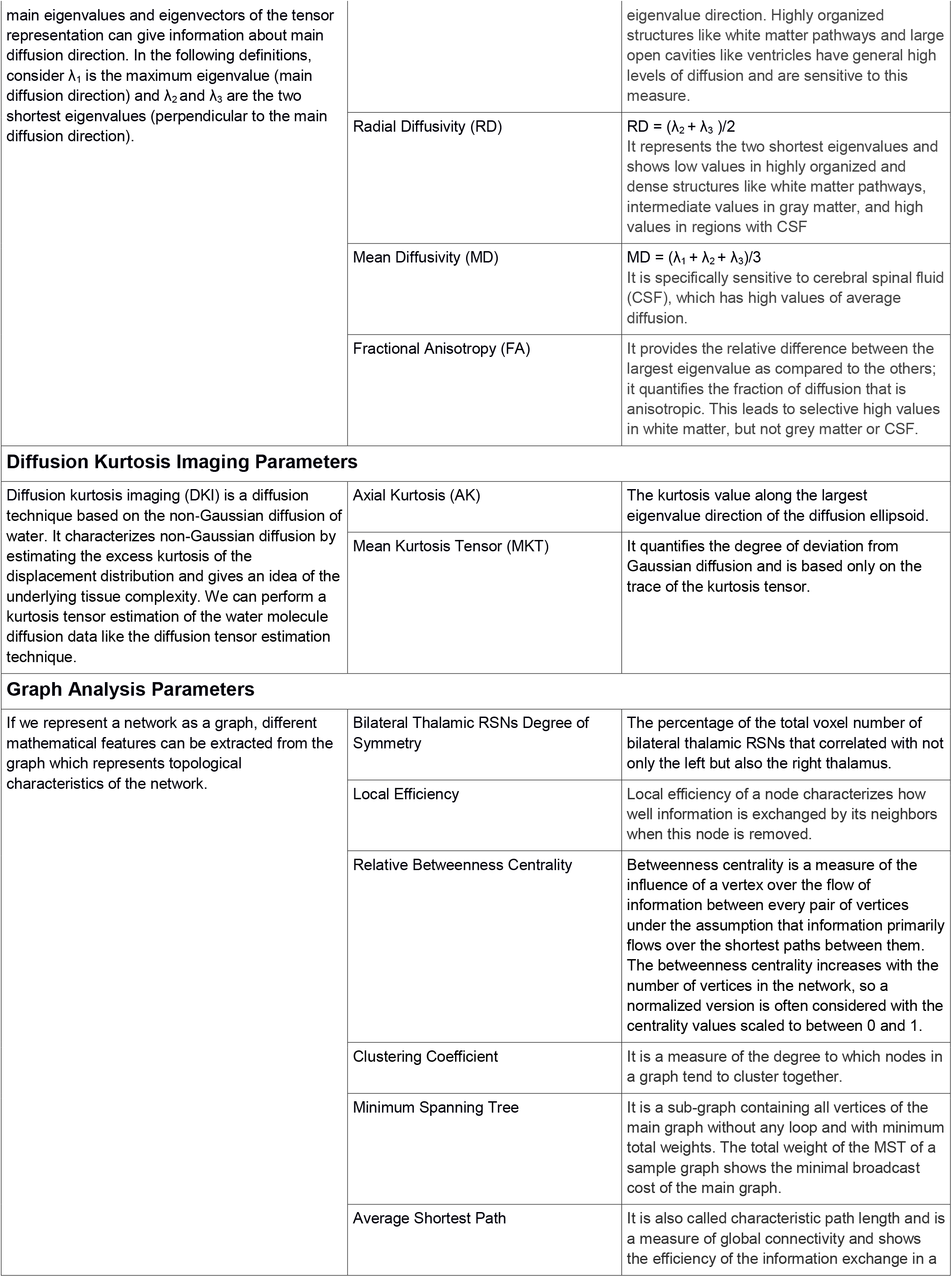
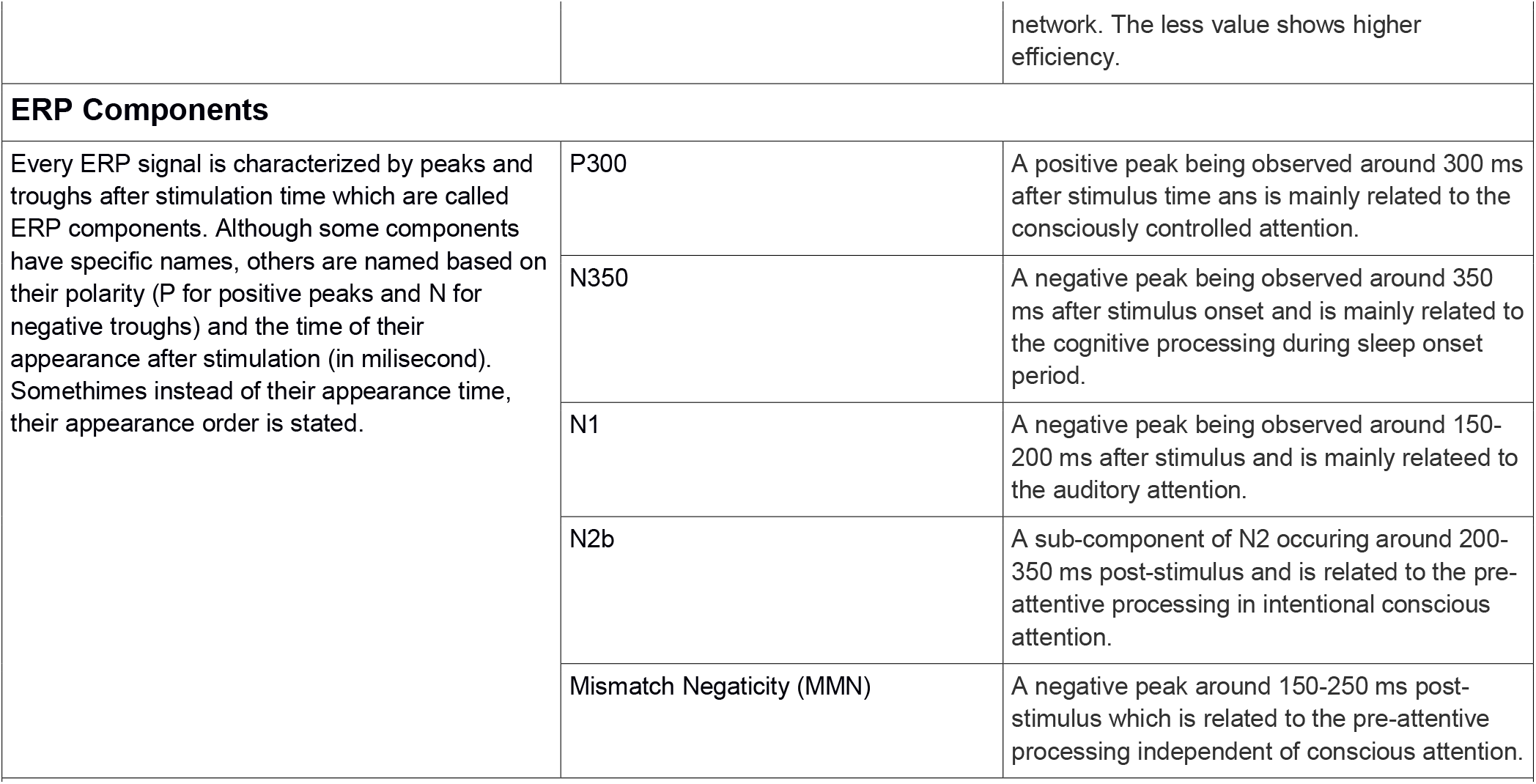
Definition of some specific outcome measures in the reviewed papers.

### 3.1 Conventional MRI

Among 12 studies retrieved, 5 showed positive results (42% specificity). The results consist of smaller volume of cingulate gyrus isthmus, ventromedial prefrontal cortex (vmPFC) and fusiform gyrus, lower thickness of frontal, parietal, and temporal regions, and higher number of intracranial bleeds (ICB) in PCS patients. MRI was acquired from total of 539 subjects, 181 of whom were for those studies with positive results (34% subject specificity).

### 3.2 Diffusion Weighted Imaging

Among 27 studies retrieved, 15 showed positive results (56% specificity). The results consist of alterations of anisotropy and diffusion measures mainly in corpus callosum, longitudinal fasciculi, and tracts in the internal capsule. Data were acquired from total of 1369 subjects, 739 of whom were for those studies with positive results (54% subject specificity).

### 3.3 Functional MRI

Among 25 studies retrieved, 21 showed positive results (84% specificity). The results consist of alterations of anti-correlation which exists between default mode network and task positive network in normal conditions, and abnormal increase of thalamo-cortical connectivity (See Fig. 3). These functional data were acquired from total of 876 subjects, 773 of whom were for those studies with positive results (88% subject specificity).

**Figure 3.**
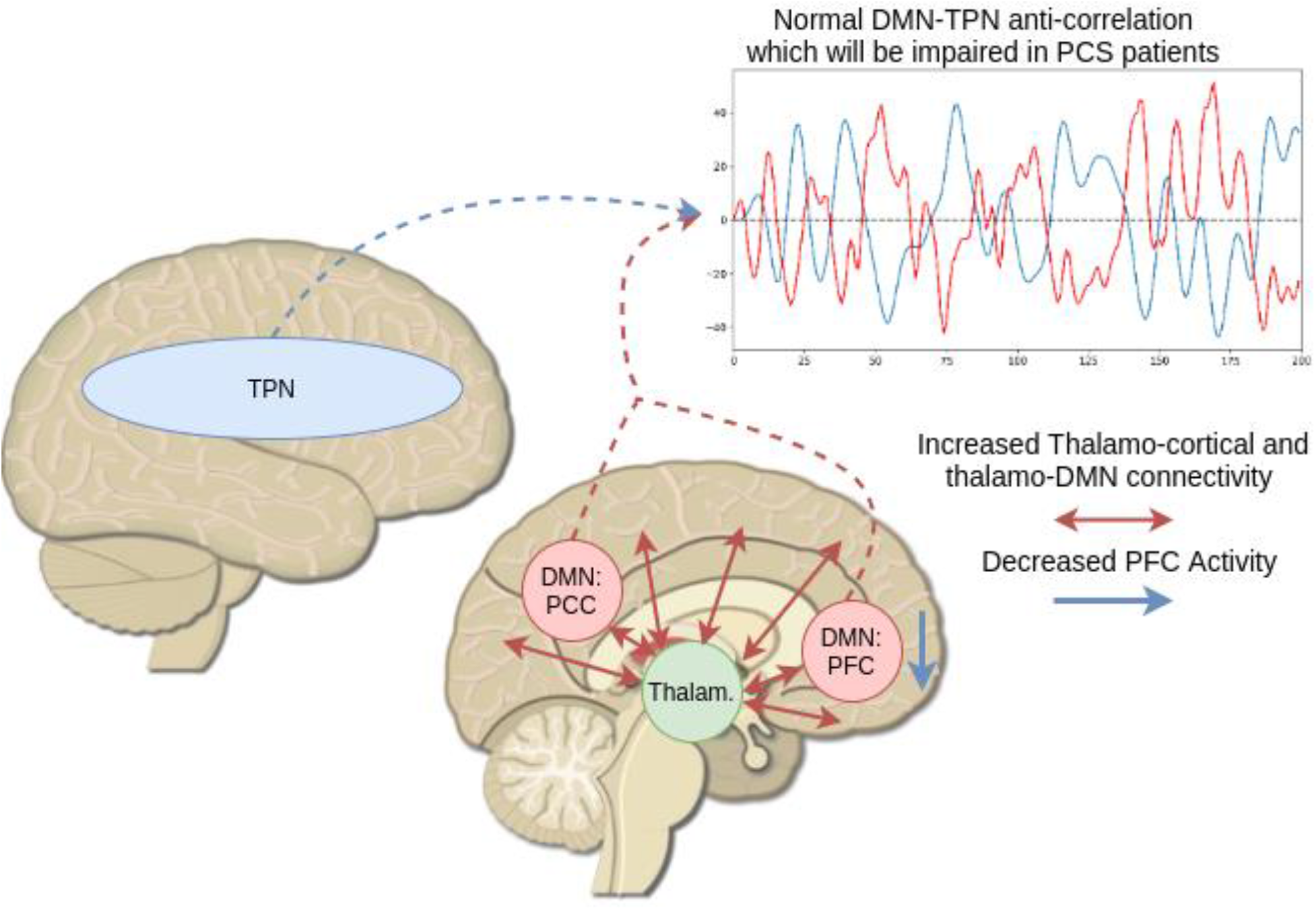
Main Findings of fMRI Studies: Based on the studies used fMRI to explore PCS, the main findings can be summarized as the abnormal increase of thalamo-cortical connectivity, alterations in the anticorrelation of default mode network and task positive network, and decrease of activity in the prefrontal cortex.

### 3.4 Electro(Magneto)encephalography

Among 10 studies retrieved (6 EEG, 4 MEG), 8 showed positive results (4 EEG, 4 MEG; 80% specificity). The results consist of reduction of ERP components such as P300, N350, N2b, N1, and mismatch negativity, increase of relative theta power, decrease of relative alpha power, decrease of beta band inter-hemispheric coherency, and abnormal slow-wave activity in orbito-frontal cortex (OFC), vmPFC, and fusiform gyrus. These data were acquired from total of 425 subjects, 375 of whom were for those studies with positive results (88% subject specificity).

## 4. Discussion

This article aims to review the current state of the art in the neuroimaging and neurophysiological aid to the diagnosis of PCS. Out of 88 papers studied in this scoping review, most of them were about structural/functional MRI and EEG/MEG. Most positive findings were found among studies using functional approaches, such as fMRI, EEG/MEG. However, the number of studies using electrophysiological techniques (i.e., EEG) remains limited compared to neuroimaging studies (i.e., fMRI), while the former are more practical and more available in clinical practice. The main findings per technique are discussed below respectively for conventional MRI, DWI, fMRI, and EEG/MEG.

### 4.1 Conventional MRI and DWI

Considering both conventional MRI and DWI studies, only 51% of the structural MRI studies [920 subjects (48% of the total subjects)] identified specific abnormalities in patients with PCS. Although DWI studies showed more specificity compared to the conventional MRI (56% vs 42%), they still have a low specificity compared to functional MRI acquisitions (56% vs 84%). Structural MRI allows to evaluate anatomical and structural brain damage in a qualitative or quantitative way. Based on specific sequence that can be used for data acquisition, different anatomical features can be studied. For example, a conventional MRI sequence is suitable to quantify and analyse volume and thickness changes of different brain regions. It is known that brain volume loss (whether globally or regionally) is a consequence of moderate or severe brain injuries (Zhou et al. 2013). For example, anterior cingulate and left cingulate gyrus isthmus in the white matter are among the regions which show atrophy over time after injury (Zhou et al. 2013). Because anterior parts of the cingulum play vital roles in different cognitive systems (e.g. working memory; Egner and Hirsch 2005), atrophy in this area can explain different persistent symptoms after the concussion. Grey matter volume within vmPFC and the fusiform gyrus was also found to positively correlate with PCS symptoms, specifically emotional functioning (Killgore et al. 2016). This is due to the fact that vmPFC has a strong connection with amygdala and involves in self-control and decision making (Motzkin et al. 2015). Furthermore, cortical thickness in frontal, parietal, and temporal regions is found to be inversely associated with self-reported symptoms in young adults with PCS (Albaugh et al. 2015). This widespread effect was linked to the repetitive blows to the head specially for the athletes in full contact sports. Besides studying changes of the volume or thickness of different brain regions, intracranial bleeds can also be analysed using susceptibility-weighted sequences. ICBs usually can be missed in CT, conventional MRI, or T2* weighted images (Liu et al. 2014). Using susceptibility-weighted images, it was shown that the number and size of the intracranial bleeds are correlated with PCS severity (Liu et al. 2014).

A widely used MRI sequence to study structural changes in the brain of PCS patients is DWI which is mainly used to study changes of the white matter tracts. To analyse these tracts quantitatively, four main parameters of fractional anisotropy, axial diffusivity, radial diffusivity, and mean diffusivity are mainly explored. FA shows the degree of anisotropy of water molecules in the white matter which ranges from 0 (isotropic movement) to 1 (anisotropic movements). Alterations in FA can infer alterations in the axonal diameter, fiber density or myelin structure. On the other hand, diffusion parameters give a value of water molecule diffusion along the main tract (AD), perpendicular to the main tract (RD), or in average (MD). PCS-associated white matter abnormalities can mainly be characterized by decrease in FA values and increase in MD, AD, and RD values in the injured tracts (FA and MD were the most reported parameters to be altered compared to others). The main reported tracts with axonal injury in the PCS patients were Corpus Callosum (Smits et al. 2011, Messé et al. 2011, Messé et al. 2012, Yeh et al. 2014, and Lange et al. 2015, Yin et al. 2019), longitudinal fasciculi (Smits et al. 2011, Messé et al. 2011, Messé et al. 2012, Lange et al. 2015, Alhilali et al. 2015, and Yeh et al. 2017), and tracts in the internal capsule (Smits et al. 2011, Messé et al. 2012, Yeh et al. 2014, Lange et al. 2015, and Alhilali et al. 2015, Yin et al. 2019). As these tracts are the main pathways of a widespread cortical and subcortical connections, any axonal damage in these tracts can lead to various cognitive symptoms in patients with PCS. These axonal damages are mainly due to the rotational acceleration and/or deceleration forces happening when the concussion occurs (Smits et al. 2011). In addition to these main tracts, altered measures of anisotropy and diffusivity were also reported in fronto-occipital fasciculi (Smits et al. 2011, Messé et al. 2011, Yin et al. 2019), uncinate fasciculus (Smits et al. 2011, Thomas et al. 2017, Yeh et al. 2017), cortico-spinal tract (Messé et al. 2011), thalamic radiation (Messé et al. 2011, Yeh et al. 2014, Lange et al. 2015, Yeh et al. 2017), cingulum bundle (Yeh et al. 2014), pontine tegmentum (Delano-Wood et al. 2015), and primary and association cortices (Fakhrin et al. 2013). As we can see and also highlighted by (Miller et al. 2016), axonal damages in patients with PCS are spatially heterogeneous and divers which could be due to the heterogeneity of the studied populations (athletes versus normal people versus veterans) and the heterogeneity of the symptoms (e.g., stress, headache, sleep disturbances, attentional issues). By applying tractography algorithms on the DWI images, structural connectivity can also be analysed between different cortical regions. Using this technique, it has been shown that, compared to the PTSD-only (post-traumatic stress disorder) patients, subjects with PCS and PTSD have greater diversity in their structural connectivity between the hippocampus and striatum, which could be associated with PCS but not PTSD (Rangaprakash et al. 2017).

In summary, the structural neuroimaging studies evaluating grey matter and white matter integrity found impairments of the cingulate gyrus and the prefrontal area, while for the white matter the corpus callosum, the longitudinal fasciculi, and internal capsule were the most frequently reported structures to be impaired or linked to PCS severity. In general, diverse structural sites have been reported to be affected by concussion and lead to PCS. This could be due to the heterogeneity of the studied populations (athletes versus regular people versus veterans) and the heterogeneity of the symptoms (e.g., stress, headache, sleep disturbances, attentional issues).

### 4.2 Functional MRI

In fMRI studies, results can be presented based on changes in BOLD activation level or based on functional connectivity among regions (i.e., network-based analysis). Furthermore, functional networks can be summarized by graph analysis metrics which create a mathematical framework to study brain networks topology in normal and pathological states in more details. We discuss fMRI findings of PCS under these different categories.

#### 4.2.1 BOLD Activation Level Analysis

Among the studies using this technique, most of them looked at working memory and selective attention deficits which are the two main cognitive domains affected after mTBI (Smits et al. 2009). Based on previous studies, brain areas which are involved in these cognitive domains are mainly dorsolateral and ventrolateral prefrontal cortex, supplementary motor area, premotor area, posterior parietal area and anterior cingulate cortex (Egner and Hirsch 2005; Owen et al. 2005; Smits et al. 2009). Activation level of these areas during tasks requiring a high cognitive load are correlated with PCS severity showing increased attentional and short-term memory demands (Pardini et al. 2010; Smits et al. 2009). Some studies showed a hyper activation also in parahippocampal gyrus and posterior cingulate cortex (PCC) which are mainly related to episodic memory and memory retrieval (Smits et al. 2009). Activation of these regions in high-load working memory and selective attention tasks shows a potential cerebral compensatory response to possible microstructural injury in patients with PCS (Smits et al. 2009). Hyperactivity was also observed in the left angular gyrus which resulted in the same behavioural accuracy for patients with PCS and healthy subjects in performing a working memory task (Gosselin et al. 2012). Taking all together, it can be stated that patients with PCS use more cognitive resources to perform an attention or working memory task with the same accuracy as the healthy subjects (Pardini et al. 2010).

Decrease in BOLD activation level in the medial prefrontal cortex was also observed in patients with PCS (Gosselin et al. 2011; Van der Horn et al. 2016). We know that this region is important for executive functioning and emotion regulation (Euston, Gruber, and McNaughton 2012; Van Der Werf et al. 2000). In addition, it is a core area of the default mode network and an important relay station between DMN and other executive networks (Van der Horn et al. 2016). As a result, impairment in this area can lead to cognitive and affective impairments (Gosselin et al. 2011; Van der Horn et al. 2016). Using transcranial magnetic stimulation as a treatment, an increase of the medial prefrontal cortex activation level during a working memory task was found and was associated with less severe PCS symptoms (Koski et al. 2015).

Besides working memory and selective attention, light sensitivity and headaches were also studied using a visual tracking task (Astafiev et al. 2016). Higher BOLD activity in the middle temporal/lateral occipital regions was observed compared to healthy subjects when patients were exposed to visual stimuli. These regions are known as extra-striate visual regions involved in motion and object processing (Astafiev et al. 2016). Higher activation in these regions was also associated with higher FA values in the same region (Astafiev et al. 2016). In sum, the light sensitivity and associated headaches could be related to an abnormal sensitivity of motion sensitive neurons.

#### 4.2.2 Functional Connectivity Analysis

In a general view, abnormal functional connectivity can be found in almost all the functional networks which can be detected in the resting state (Stevens et al. 2012). While few studies have reported an abnormal whole brain hyper-connectivity especially in acute phase (Kaushal et al. 2019), other studies have shown an overall whole-brain connectivity reduction in these patients (Palacios et al. 2017; Sours, Chen, et al. 2015). Whole-brain connectivity analysis gives us a general view of the functional organization of the brain; however, focusing on specific networks or regions, chosen in a hypothesis driven way, gives more insights about network configuration changes and more fine-grained understanding of the pathology of PCS. Among different networks and regions, DMN and the thalamus are the most studied ones.

##### Default Mode Network – DMN

The DMN is a network mainly related to the internal awareness (Vanhaudenhuyse et al. 2011). Posterior regions of the DMN, such as the precuneus, showed more vulnerability to traumatic injury (Zhou et al. 2012). Decrease of the connectivity of the precuneus inside the DMN and its increased connectivity with primary and secondary visual networks and left frontoparietal network can alter the transition of attention from internal to external awareness (Zhou et al. 2012). Another critical area in the DMN is the PCC. Decreased connectivity of the PCC within the DMN is associated with increased connectivity of medial prefrontal cortex (MPFC), which potentially shows the compensatory role of the MPFC to the impaired neurocognitive functions. Indeed, the MPFC and the PCC are intrinsically interdependent inside the DMN and their function is highly complementary (Zhou et al. 2012). Anterior regions of the DMN such as the anterior cingulate cortex (ACC) have also shown altered connectivity in these patients. Connectivity of the ACC with emotional, primary visual, and higher order processing networks is negatively correlated with the severity of the PCS (Stevens et al. 2012). Considering the role of the ACC in pain processing, error and conflict detection, behavioural and cognitive control, as well as emotional processing, this could potentially explain different complaints in mTBI patients. In addition to the role of the DMN in internal awareness, the interaction between the DMN and the task positive network (TPN) is crucial for performing executive functions. In fact, the balance between internal and external awareness is supported by the interaction of these two networks with temporally alternating higher activity states (Di and Biswal 2014). In patients with PCS, the deactivation of the DMN is not strong enough while they are performing a working memory task (Van der Horn et al. 2016). Specifically, an increase in the connectivity between the DMN and the TPN has been observed (Van der Horn et al. 2016; Sours, George, et al. 2015, Manning et al. 2019). This altered anti-correlation between the DMN and the TPN (i.e. the lack of silencing of the DMN) could explain some of the difficulties in task performance that patients with PCS suffer from.

##### Thalamus

This subcortical hub plays an important role in memory, executive functions, attention, and emotion, among others (Van Der Werf et al. 2000). Different studies have shown a PCS related increase of the thalamo-cortical connectivity (Sours, George, et al. 2015; Tang et al. 2011; Zhou 2017) which can lead to an inefficient neuronal activity (Zhou 2017). The increased thalamo-cortical connectivity can be related to the alteration of the GABAergic inhibitory neurons in the thalamus (Sours, George, et al. 2015; Tang et al. 2011), resulting in a reduction of inhibitory control over the thalamocortical activity. Connectivity of the thalamus with other networks has also been studied. Especially, evidence suggests that the connectivity between the thalamus and the DMN increases in patients with PCS (Banks et al. 2016; Sours, George, et al. 2015). As mentioned, well-coordinated interaction between the DMN and the TPN is necessary for an appropriate balance between internal and external awareness and proper task execution. Fronto-parietal control (FPC) network (Banks et al. 2016) and salience network (SN) (Seeley et al. 2007) were suggested as networks that control this balance between the DMN and the TPN. The thalamus has connections with all of these networks and has a key role in this complex interaction between networks. Increased connectivity between the thalamus and the DMN could lead to a malfunctioning of this modulatory interaction. In addition to the increased connectivity of both thalamus and DMN, a decrease of connectivity between the thalamus, the FPC and the DAN was also reported (Banks et al. 2016). A normalization of this decreased connectivity was correlated with better PCS scores, while the connectivity with the DMN remained unchanged.

##### Other Regions

Besides DMN and thalamus, hypo-connectivity of some anterior regions such as inferior and middle frontal gyri was also reported in PCS patients (Murdaugh et al. 2018). Inferior and middle frontal gyri are part of the active cognition network which should be anti-correlated with the DMN. Their abnormal connectivity disrupts the balance that should exist between the DMN and executive networks. Reduction of homotopic functional connectivity in pain processing regions is another finding which was shown to be normalized with symptom recovery and can be serve as another potential biomarker of PCS (Chong et al. 2019).

#### 4.2.3 Graph Analysis

One approach to study brain networks and their topological characteristics is, the data driven, graph analysis. In this approach, connectivity measures are represented by weighted graphs (nodes represent regions of interest or ROIs and edges represent connectivity between ROIs). For example, graph modularity (i.e. the strength of division of brain network into modules) is increased in the temporal regions in the subacute phase and are decreased in frontal regions in the chronic stage in patients with PCS (Messé et al. 2013). These changes can be localized mainly in regions related to executive functions, working memory, cognitive control and attention. Some mathematical features of the graph (detailed in Table 2) such as the clustering coefficient is also correlated with increased thalamo-cortical connectivity (Zhou 2017). Combining graph analysis results of the functional connectivity data with the diffusion weighted images revealed that regions of the brain showing lower local relative betweenness centrality in the functional network have also lower fractional anisotropy; in addition, decrease of global efficiency of functional networks is related to diffuse axonal injury (Zhou 2017). Average shortest path weights and minimum spanning tree weights have also been shown to be increased in patients with PCS (Hou et al. 2019). Taking all these results together, graph analysis shows that patients with PCS tend to have costly and less efficient network configurations for information transfer (Hou et al. 2019) which leads to cognitive impairments.

To summarize, findings of 84% papers which have reported significant results (with a total of 773 subjects) showed that impairment in functional connectivity after mTBI could potentially explain most of the post-concussive symptoms. Mainly, alterations of the connectivity in the DMN and its interaction with other executive networks, especially alterations of the anti-correlation between the DMN and the TPN, could be associated with cognitive impairments. In addition, increase of the thalamo-cortical connectivity due to alteration of GABAergic inhibitory neurons in the thalamus can also lead to difficulties in task performance. These functional impairments recruit compensatory mechanisms in the brain and use of extra cognitive resources while performing cognitive tasks.

### 4.3 Electro(Magneto)encephalography

In general, EEG/MEG data can be analysed in the sensor space (signals directly from surface sensors) or in the source space (i.e. signals of surface sensors are projected to the cortex to estimate the cortical location of the neural generators). Furthermore, different analysis methods can be performed on the standard frequency bands of the signals: delta (0-4 Hz), theta (4-8 Hz), alpha (8-12 Hz), beta (12-30 Hz), and gamma (>30 Hz). In addition, EEG/MEG recordings can be either in resting state or in response to stimuli, averaged over many trials, to measure event related potentials (ERP) in case of EEG recording or event related fields (ERF) in case of MEG recording. In ERP/ERF signals, the amplitude and latency of their different components are important and meaningful features to study. Although in the analysis methods, there are no serious differences between EEG and MEG signals, in terms of clinical applicability and data acquisition, they are completely different. While EEG is widely available and can be performed easily in different clinical settings, MEG is scarcely available for clinical purposes. In the present review, 8 out of 10 EEG/MEG studies identified neurophysiological changes linked to PCS (6 EEG studies with 4 reporting positive results and 4 MEG studies all reporting positive results).

Considering ERP studies, they were mainly focused on the amplitude of P300 (Gosselin et al. 2012, Ruiter et al. 2019), N350 (Gosselin et al. 2011, 2012), N2b, N1, and mismatch negativity (MMN) components (Ruiter et al. 2019). All of them reported a reduction in the amplitude of such components. A first ERP study found that amplitude of N350 component (mainly related to cognitive processing during sleep onset period (Harsh, et al. 1994)) in frontal electrodes was decreased in patients with PCS while performing a working memory task compared to a control task (Gosselin et al. 2011, 2012). Because this N350 amplitude reduction was correlated to the lower BOLD signal changes of the right mid-dorsolateral prefrontal cortex (mid-DLPFC) in the same task, it is thought that this specific ERP component is related to the neural activity of the mid-DLPFC. In addition, the amplitude of P300 (mainly related to consciously controlled attention) in parietal regions is decreased after brain injury and this reduction in amplitude is more significant when patients with PCS suffer from severe depressive symptoms (Gosselin et al. 2012). While both N2b and MMN are related to the pre-attentive processing, MMN is observed independent of the conscious attention and N2b is observed in intentional conscious attentions (Ruiter et al. 2019). As a result, reduction of amplitude of P300, N2b and MMN can be linked to the attentional deficits which are common in PCS. The N1 component mainly reflects auditory attention and shows a reduced amplitude in case of auditory processing difficulties (Ruiter et al. 2019). At the network level, brain network activation (BNA) is a task related EEG feature (Shahaf et al. 2012) that could act as a reliable biomarker of diagnosis and long-term tracking of brain networks after concussion (Reches et al. 2017). This parameter is indeed decreased in patients with PCS compared to healthy subjects immediately after injury in association with symptom severity, and may be linked to a reduced task-related brain activity and connectivity in these patients (Reches et al. 2017). In analysis of resting state EEG data, increase of relative theta power, decrease of relative alpha power and decrease of beta band inter-hemispheric coherence have been observed (Lewine et al. 2019).

Considering MEG recordings, it has been shown that total MEG magnitude in the source space is correlated with PCS scores (Huang et al. 2014). Still, slow-wave features are more informative for cognitive impairment examination in this cohort of patients (Robb Swan et al. 2015). For example, abnormal slow-wave activity in the orbitofrontal cortex and vmPFC has been shown to be associated with personality changes, concentration troubles, and emotional instability (Huang et al. 2014). Because these regions are highly connected to other cortical and subcortical areas, this abnormal activity can cause impairments in a vast domain of cognitive abilities (Huang et al. 2014). In addition, visual difficulties such as blurred vision have been explained by the abnormal slow-wave activity in the right fusiform gyrus (Huang et al. 2014), an important region for face, object, and body recognition and processing (Weiner and Grill-Spector 2010). Abnormal slow-wave generation in different regions has been associated with white matter abnormalities or micro-structure damages to the major tract in the same regions (Huang et al. 2009). Connectivity analysis features of MEG data in different frequency bands have also been shown to be informative biomarkers to study PCS after mTBI. Inter-regional resting-state phase synchrony in the alpha band could classify patients from healthy subjects with an accuracy of 80%, in which the distance of each participant to the classification boundary was correlated with the symptom severity of that subject (Vakorin et al. 2016). Functional connectivity of ROIs in prefrontal cortex, medial temporal lobe, putamen, and cerebellum with the whole brain was increased while functional connectivity of the right frontal pole with the rest of the brain in patients with PCS was decreased (Huang et al. 2017). These abnormal connectivity variations were observed in beta, gamma, and low frequency bands (i.e. 1-7 Hz). Decrease of the functional connectivity in the right frontal pole can be explained by diffuse axonal injury which leads to disruption of neural communications (Hannawi and Stevens 2016). On the other hand, the increase of functional connectivity in prefrontal cortex, medial temporal lobe, putamen, and cerebellum can be explained by the over excitation of glutamate in pyramidal neurons of the cortex which leads to an alteration of GABAergic interneurons (Huang et al. 2017).

Taking all the information together, different alterations in the EEG/MEG markers may be related to the white matter abnormalities and axonal injury, alterations of GABAergic neurons and functional dysfunction of some frontal regions such as the DLPFC.

### 4.4 Limitations

Our work had also some limitations. First of all, heterogeneity of studies population resulted in heterogeneity of symptoms and imaging markers. Secondly, few number of studies explored PCS longitudinally. These two limitations made us unable to have specific strong prognostic recommendations for PCS. Third limitation was the few number of studies used other imaging techniques than MRI and EEG. While imaging techniques like PET, SPECT, and fNIRS can give us more insight about the brain alterations in PCS, the few number of studies limited generalization of their findings and made us exclude them from our analysis. Finally, most of the studies we included in our survey didn’t have a specific hypothesis and the questions they were trying to answer were really divers. Therefore, we couldn’t perform a meta-analysis which could help us to understand PCS more deeply.

## 5. Concluding remarks

In this scoping review, we studied the most reliable approaches to evaluate PCS and the common findings among studies which could explain PCS (i.e., biomarkers of PCS). Most of the studies using structural MRI to investigate neural correlates of PCS did not find any specificities, except for an atrophy mainly involving the cingulate and prefrontal areas linked to PCS symptoms, and abnormalities in the anisotropy and diffusion parameters of corpus callosum, longitudinal fasciculi, and internal capsule. On the other hand, the majority of functional studies (fMRI and EEG/MEG) found abnormal increase in thalamo-cortical functional connectivity, which could be linked to employment of compensatory mechanisms needed to perform cognitive tasks. In addition, an altered anti-correlation between the DMN and the TPN could explain some of the difficulties in task performance experienced by patients with PCS. Besides, reduction of brain activity in specific areas was also found; the most frequently reported ones being the prefrontal cortex. Functional abnormalities in these abovementioned brain regions and networks, should draw physicians’ attention to provide appropriate cares to these at-risk patients. Note that, only six studies used EEG, although compared to MRI, or even MEG, this technique is easier to use in clinical practice. Further EEG studies could provide insightful prediction of PCS outcome that could be more easily implemented in clinical practice. EEG/ERP studies identified a reduction in ERP component amplitudes related to attentional processes. It should be noted that in most of the studies, no a priori or precise hypotheses were stated. We hope that our findings will guide authors to postulate specific hypotheses for future neuroimaging studies. So far, fMRI seems the most robust approach to study PCS severity and future prospective longitudinal studies in a large sample of concussed patients using clinically relevant techniques such as EEG, should determine prognostic factors of good and poor outcome following a concussion.

## Data Availability

Not applicable

## Acknowledgement

The study was further supported by:

The study was supported by the University and University Hospital of Liege, the Belgian National Funds for Scientific Research (FRS-FNRS), the European Union’s Horizon 2020 Framework Programme for Research and Innovation under the Specific Grant Agreement No. 945539 (Human Brain Project SGA3) and No. 686764 (Luminous project), DOCMA project [EU-H2020-MSCA– RISE–778234], the European Space Agency (ESA) and the Belgian Federal Science Policy Office (BELSPO) in the framework of the PRODEX Programme, “Fondazione Europea di Ricerca Biomedica”, the Bial Foundation, the Mind Science Foundation and the European Commission, the fund Generet, the Mind Care International Foundation and the King Baudouin Foundation.

The authors have no conflict of interest relevant to the present study to declare.

## List of abbreviations

ACC: anterior cingulate cortex
AD: axial diffusitivity
AI: axonal injury
BC-PSI: British Columbia postconcussion symptom inventory
BOLD: blood-oxygen-level dependent
CT: computed tomography
DAN: dorsal attentional network
DMN: default mode network
DLPFC: dorsolateral prefrontal cortex
DSM-IV: diagnostic and statistical manual for mental disorders – IVth edition
DTI: diffusion tensor imaging
DWI: diffusion weighted imaging
EEG: electroencephalography
ERF: event related field
ERP: event related potential
FA: fractional anisotropy
FC: functional connectivity
FLAIR: fluid-attenuation inversion recovery
fMRI: functional magnetic resonance imaging
fNIRS: functional near infrared spectroscopy
FPC: fronto-parietal cortex
GM: grey matter
GO: good outcome
ICBs: intracranial bleeds
ICD-10: international classification of diseases – 10th edition
IFO: inferior fronto-occipital fasciculus
ImPACT: immediate post-concussion assessment and cognitive testing
MD: mean diffusitivity
MEG: magnetoencephalography
MKT: mean kurtosis tensor
MMN: mismatch negativity
MPFC: medial prefrontal cortex
MRS: magnetic resonance spectroscopy
mTBI: mild traumatic brain injury
OFC: orbitofrontal cortex
PCC: posterior cingulate cortex
PCD: Post-concussion disorder
PCS: post-concussion syndrome
PCS-: post-concussion syndrome negative
PCS+: post-concussion syndrome positive
PCS-NIM: PCS-negative impression management
PCSQ: post-concussive symptom question-naire
PET: positron emission tomography
PO: poor outcome
PTSD: Post-traumatic stress disorder
RD: radial diffusitivity
ROI: region of interest
RPQ: Rivermead post concussion symptoms questionnaire
RSN: resting state network
SCAT: sport concussion assessment tool
SN: salience network
SP: shortest path
SPECT: single photon emission computed tomography
SWI: susceptibility-weighted imaging
TBI: Traumatic brain injury
TPN: task positive network
vmPFC: ventromedial prefrontal cortex
WM: white matter

